# Complementary roles of structure and variant effect predictors in RyR1 clinical interpretation

**DOI:** 10.1101/2025.04.02.25325085

**Authors:** Rolando Hernández Trapero, Mihaly Badonyi, Lukas Gerasimavicius, Joseph A Marsh

## Abstract

RyR1-related disorders, arising from variants in the RYR1 gene encoding the skeletal muscle ryanodine receptor, encompass a wide range of dominant and recessive phenotypes. The extensive length of RyR1 and diverse mechanisms underlying disease variants pose significant challenges for clinical interpretation, exacerbated by the limited performance and biases of current variant effect predictors (VEPs). This study evaluates the efficacy of 70 VEPs for distinguishing pathogenic RyR1 missense variants from putatively benign variants derived from population databases. Existing VEPs show variable performance. Those trained on known clinical labels show greater classification performance, but this is likely inflated by data circularity. In contrast, VEPs using methodologies that avoid or minimise training bias show limited performance, likely reflecting difficulty in identifying gain-of-function variants. Leveraging protein structural information, we introduce Spatial Proximity to Disease Variants (SPDV), a novel metric based solely on three-dimensional clustering of pathogenic mutations. We determine ACMG/AMP PP3/BP4 classification thresholds for our method and top-performing VEPs, allowing us to assign PP3/BP4 evidence levels to all RyR1 missense variants of uncertain significance. Thus, we suggest that our protein-structure based approach represents an orthogonal strategy over existing computational tools for aiding in the diagnosis of RyR1-related diseases.

## Introduction

RyR1, encoded by the *RYR1* gene, is the skeletal muscle ryanodine receptor and the largest human ion channel, with a sequence spanning 5038 amino acids and a molecular weight of ∼2.2 MDa [1, 2]. The protein serves as a calcium release channel of the sarcoplasmic reticulum, as well as a bridging structure connecting the sarcoplasmic reticulum and transverse tubule. The monomeric subunits self-associate to form homotetrameric assemblies involved in excitation-contraction (EC) coupling. Four-fifths of the RyR1 protein is cytoplasmic and the remaining one-fifth consists of lumenal and membrane spanning domains [3]. Small molecule ligands to RyR1 include Ca^2+^, ATP, and caffeine, along with several pharmaceuticals such as volatile anaesthetics (*e.g.,* halothane), ryanodine (channel agonist), and dantrolene (channel antagonist) [4–6]. Given its complex regulation and essential role in calcium homeostasis, RyR1 is highly sensitive to disruption by genetic variation.

Consistent with this, mutations in RyR1 have been implicated in a wide and highly heterogeneous set of genetic disorders [6–8]. Pathogenic RyR1 variants occur across the sequence and may disrupt regulatory interactions, hypersensitizing the protein to lower voltage thresholds, or cause gain or loss of function through other mechanisms [6]. Malignant hyperthermia (MH) is an autosomal dominant disorder that can be explained by increased probability of channel opening due to a reduction in the threshold needed for calcium induced calcium release. This leads to leaky channels under high temperatures or exposure to triggering agents, such as volatile anaesthetics, muscle relaxants, and caffeine. Dominant variants can also cause central core disease (CCD) due either to receptor hypersensitization, resulting in calcium dysregulation and depletion of calcium stores even under resting conditions, or affecting the voltage-dependent activation of calcium release (EC uncoupling) [9–11]. Unusually, central core/rod disease (CCRD) can result from excessive production of RyR1 [12]. In contrast, recessive mutations that cause multi-minicore disease (MmD), centronuclear myopathy (CNM) and congenital fiber-type disproportion (CFTD) reduce RyR1 expression or activity, *i.e.,* cause a loss of function. This phenotypic diversity suggests that RyR1-related disorders may be best conceptualised as a spectrum, rather than as discrete diseases [13–16].

This diversity in clinical outcomes also complicates diagnosis, particularly due to the high number of variants of uncertain significance (VUS) identified in *RYR1*. The extreme length of the *RYR1* gene means that the chance of finding one or more VUS when sequencing the gene in a patient is very high. Upon identification of a VUS, a patient will often be referred for further testing. Especially in the case of malignant hyperthermia, the *in vitro* contracture test (IVCT) can be useful for diagnosis in the event of finding a VUS in a clinical panel [17]. However, this solution has its disadvantages, as it is done on a per-patient basis without the purpose of providing mechanistic insight. Furthermore, a negative IVCT does not preclude the occurrence of a different but related RyR1 dysfunction. Thus, if computational approaches could be used to better exclude VUS that have little chance of having clinically meaningful effects, or prioritise those with a high likelihood of pathogenicity, this could have a significant benefit in terms of improving diagnosis and reducing the burden of patient testing. This has motivated growing interest in computational methods for variant interpretation.

Variant effect predictors (VEPs) are tools designed to score the likely damaging effects or pathogenicity of genetic variants [18]. Compared to the experimental characterization of variants, these have the advantage of being very fast and essentially free to run. However, given the large number of VEPs that have been released, it can be difficult to know what the best methods are to use. Moreover, the performance of new VEPs as reported upon publication tends to be highly overstated [19], and thus the extent to which their predictions can be relied upon is often unclear. The performance of VEPs can also vary dramatically between different genes, with their utility being markedly worse for genes associated with dominant-negative and gain-of-function molecular mechanisms [20]. Given that the extensive experimental evidence for gain-of-function effects in RyR1 [9, 10, 21–23] along with recent computational predictions showing that, overall, pathogenic RyR1 variants were most likely to be associated with gain-of-function effects [24], this suggests that current VEPs might struggle with them.

In this study, we have performed an analysis of known missense variants in RyR1, with the goal of improving our ability to computationally predict their effects. First, we evaluate currently available VEPs for their ability to distinguish between pathogenic and putatively benign variants. While certain clinical-trained methods appear to perform well, we find that this is likely due to data circularity that arises from these methods having been trained using known RyR1 variants. In contrast, population-free VEPs, which are free from this bias, show substantially worse performance. Next, we investigate the protein structural context of RyR1 missense variants. While we find that the effects of variants on protein stability have little power to explain pathogenicity, consistent with a likely gain-of-function disease mechanism, we observe strong clustering of pathogenic missense variants within three-dimensional space. Based upon this, we introduce a new method, based solely on spatial proximity to known pathogenic mutations, which we show performs comparably to the best population-free VEPs for the identification of pathogenic missense variants in RyR1. Thus, our structure-based approach holds complementary value to existing computational tools for aiding the diagnosis of RyR1-related disorders.

## Methods

### Variant effect predictor scores and analysis

To construct an aggregated set of variant effect predictor results, we used an in-house pipeline to provide these scores for every possible missense variant in RyR1 [25]. In total, 70 variant effect predictors were used in the analyses. For DeepSequence [26], due to the size of the RyR1 protein, it was necessary to split the protein into 7 segments of 800 amino acids (538 for the final segment) with a 50 amino acid overlap. We used Jackhmmer to produce a multiple sequence alignment for each segment from the UniRef100 database with an inclusion threshold of 0.5 bits per residue. DeepSequence was run locally on a GPU for each segment individually to obtain prediction scores for all variants, and mean scores were taken for the overlapping regions.

For the evaluation of variant effect predictors, we used RStudio with R version 4.4, along with tidyverse for pre-processing, ROCR for ROC AUC analyses, bio3D and spatstat for the calculation of the spatial clustering features, and ggplot and ggpubr for generating the plots. For these analyses, we removed missing values, duplicates, and made mutually exclusive pathogenic and putatively benign sets.

### Spatial clustering feature calculation

The Extent of Disease Clustering (EDC) metric was calculated as previously described [20]. The various SPDV (Spatial Proximity to Disease Variants) distance metrics were derived in R using the ‘bio3d’ package [27] to handle PDB structural data and derive distance matrices. Using residue alpha carbon atoms as the reference point, we calculated Euclidean distances from a residue of interest to all known disease variant positions in a structure. When a position of interest itself contained disease variants it was excluded from SPDV calculation, deriving the shortest distances to all other disease-harbouring positions. The distances were ranked in ascending order and a single (SPDV_1) or a number of closest distances (SPDV_2, SPDV_3, etc.) were retrieved and averaged for a given SPDV metric. Calculations were carried out using the tetrameric rabbit RyR1 complex structure 7M6A [28], which has a 97% sequence identity to human. All but two pathogenic missense variants occur at positions conserved between human and rabbit, while D3501Y and A4329D are at residues not present (*i.e.* disordered) in the 7M6A structure. A pairwise alignment of the human sequence to the PDB structure is provided as Table S1.

Importantly, SPDV is calculated using Cα–Cα distances, which are more reliably resolved than side-chain positions at moderate resolution. The RyR1 structure used has a resolution of 3.36 Å, which is sufficient for accurate backbone placement. Relying on backbone atoms reduces noise and enhances robustness, especially for large complexes where side-chain flexibility may obscure structural clustering.

For easy access, we implemented the SPDV calculation pipeline as a Colab notebook (https://edin.ac/40yke9l), which supports both monomeric and protein complex structures and allows inter-chain SPDV calculations.

### FoldX free-energy calculations

We used FoldX [29] to predict changes in the Gibbs free energy (ΔΔG) between wild-type and mutant structures, considering both intra- and intermolecular effects calculated from the full tetrameric complex (PDB ID: 7M6A) [28]. The ‘RepairPDB’ function was run before modelling, followed by ‘BuildModel’ with 10 replicates for ΔΔG calculation. We considered absolute (|ΔΔG|) values from the full protein complex structure as these tend to show the largest differences between pathogenic and putatively benign mutations [20, 30]. Rank-normalised ΔΔG_rank_ values were calculated as previously described [31] and used for plotting to aid in visualisation.

### ACMG/AMP PP3/BP4 classification of variants

The American College of Medical Genetics and Genomics (ACMG) and the Association for Molecular Pathology (AMP) recommend the use of computational predictors as supporting evidence for assessing the pathogenicity or benignity of genetic variants. To classify RYR1 missense variants under the PP3/BP4 criteria, we used the acmgscaler R package [32], which is available at https://github.com/badonyi/acmgscaler. The method calculates positive likelihood ratios using a bootstrap-based Gaussian kernel density estimation procedure, whereby each score is assigned a likelihood ratio that determines its classification according to ACMG/AMP evidence thresholds. ClinVar-assigned pathogenic or likely pathogenic (P/LP) and benign or likely benign (B/LB) variants were used as reference distributions. We adopted the 10% prior probability of pathogenicity previously proposed [33], which is also the default for *acmgscaler*. This offers a pragmatic compromise for genes like *RYR1* with mixed inheritance patterns and high VUS rates, where the true prior is difficult to estimate and likely varies depending on clinical context.

## Results and Discussion

### Compilation of known RyR1 missense variants

To obtain a set of pathogenic RyR1 missense variants to use for our analyses, we compiled disease-associated missense variants from the ClinVar, OMIM, and dbSNP databases, as well as performing an extensive search of the literature for other reported cases. In total we identified 430 missense variants that have been reported to be associated with RyR1 disease, which are listed in **Table S2** and **Table S3**. This approach gives far more disease-associated RyR1 variants than the 85 pathogenic and likely pathogenic missense variants classified with strict criteria used by the ClinGen Variant Curation Expert Panel (VCEP) [34]. While we expect that there will inevitably be some false positive cases in our disease-associated variant dataset, given the less stringent selection criteria compared to the VCEP, we think that the increased statistical power afforded by the much larger dataset is worth the trade-off.

Within our set of disease-associated missense variants, we were able to further classify these based upon clinical phenotypes: 165 associated with autosomal dominant malignant hyperthermia (MH), 71 with autosomal dominant central core disease (CCD), 123 with other autosomal dominant RYR1-related disease (RRD), and 71 with autosomal recessive RYR1-related disease. The MH and CCD groups were strictly curated to ensure no overlap in phenotypic annotations based on ClinVar entries and supporting literature. In contrast, the RRD group was introduced to account for dominant phenotypes with greater clinical heterogeneity, including variants annotated as congenital myopathy 1A, King-Denborough syndrome, or with multiple overlapping disease terms. This grouping strategy reflects the phenotypic variability often encountered in clinical practice and avoids over-fragmentation of small variant subsets. Recessive RyR1-related disease variants were also treated separately to investigate differences that might arise between this group and dominantly inherited disease variants. The distribution of these four classes of missense variants along the linear sequence of the RyR1 protein is shown in **Figure 1**.

**Figure 1:**
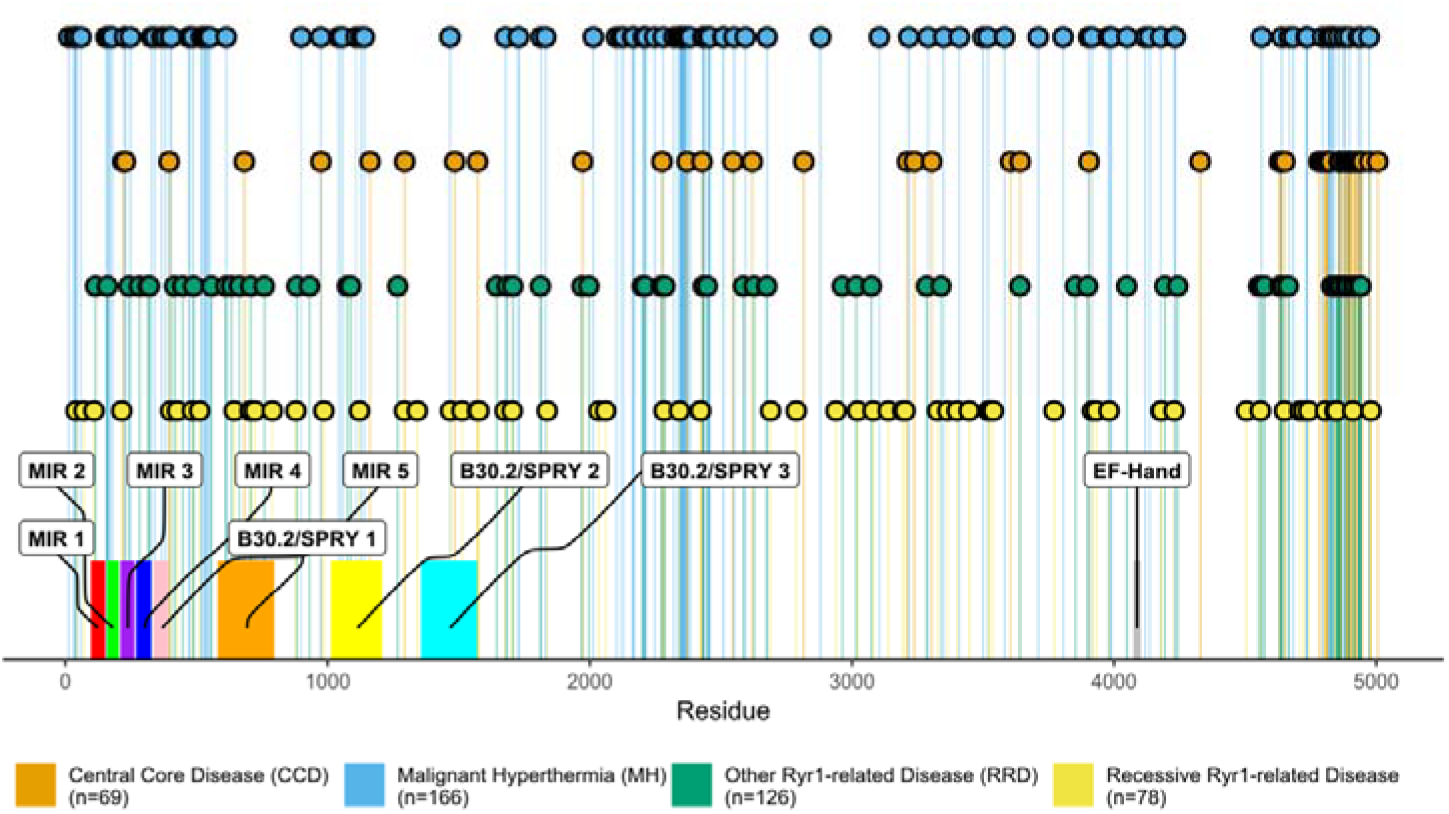
Disease-associated RyR1 mutations cluster in linear space. Lollipop plot of RYR1 variant positions by disease across the amino acid sequence with domains indicated from UniProt (ID: P21817).

For comparison, we also identified 6254 missense variants present in the human population using gnomAD v4.1, after excluding any present in our disease-associated dataset. We consider missense variants present in gnomAD at any allele frequency, excluding those from our pathogenic set, to be “putatively benign” for the purpose of analysing VEP performance, as we have done in several recent studies [35–38], as an allele frequency filter would severely reduce the number of variants in our analysis. Furthermore, it has been argued that assessing performance against rare variants present in the population is more reflective of the practical clinical utilisation of computational predictors, in which rare pathogenic variants must be distinguished from rare benign variants [39, 40]. As gnomAD is derived from a generally healthy population, we expect that the vast majority of these variants should have no clinically relevant effect, although we accept that it could contain some recessive variants in a heterozygous state, or variants associated with later onset disease or with incomplete penetrance [41].

### Evaluation of variant effect predictors for discriminating between disease-associated and putatively benign missense variants

Next, we sought to evaluate the performance of currently available VEPs at distinguishing between dominant pathogenic and putatively benign missense variants. We excluded recessive variants from this analysis, as they are frequently found in a heterozygous state in the healthy population. We therefore computed predicted effects of the available RyR1 missense variants using 70 different VEPs used in our recent benchmarking studies [40, 42]. To evaluate the performance of the VEPs as classifiers, we used receiver operating characteristic (ROC) curves and calculated their area under the curve (AUC) values. This approach is widely used for assessing binary classifiers and is robust to differences in dataset size, which is important given that not all predictors return scores for every variant.

The variant effect predictors (VEPs) can be grouped into three classes based on their exposure to human-derived data and the resulting risk of circularity [42]. *Clinical-trained* VEPs are supervised machine learning models—often using random forests or neural networks—that are explicitly trained on variants with clinical labels (*e.g.*, pathogenic and benign). As a result, they are highly susceptible to data circularity, particularly when performance is assessed on variants from the same or similar sources [43–45]. In our case, since we analyse all known *RYR1* missense variants, it is almost certain that many of these were included in the training data for clinical-trained predictors. As a result, the apparent performance of these methods on *RYR1* will likely be inflated due to overlap between training and evaluation data.

In contrast, *population-free* VEPs do not use clinical or population-derived variants during training and thus are almost completely immune from circularity concerns. These methods typically rely on features such as evolutionary conservation derived from multiple sequence alignments, or increasingly, utilise protein language models. Notably, deep mutational scanning (DMS)-based benchmarking, which provides independent ground truth, has consistently found that many of the top-performing VEPs tend to be population-free [42]. While it is possible that clinical-trained VEPs developed specifically for prioritising clinically relevant variants will have some advantages at that task compared to unsupervised methods, the greatly reduced bias of population-free VEPs means we can have much more confidence in their reported performance.

Finally, certain predictors, while not explicitly trained on clinical labels, are nonetheless tuned or calibrated using population data, such as allele frequencies. These *population-tuned* VEPs, including tools like AlphaMissense [46], exhibit reduced circularity issues compared to clinical-trained methods, but are not fully independent and may still exhibit performance inflation when tested on variants drawn from human population or clinical datasets [42, 47].

Interestingly, we observe the top 13 performing VEPs when assessing their ability to discriminate between pathogenic and putatively benign RyR1 missense variants to be clinical-trained methods (**Figure 2**). The best performance is seen for the metapredictor BayesDel_noAF (ROC AUC: 0.874) [48]. Notably, this is much better performance than observed for VARITY_R (ROC AUC: 0.789) [39], which was previously identified as the top performing clinical-trained VEP when benchmarked against independent DMS data [42], and also showed very low bias when applied to variants from different ancestry groups [47]. Given that VARITY_R was designed in a manner to minimise training bias and that BayesDel_noAF underperformed compared to VARITY_R in the benchmark, this strongly suggests that the apparent top performance of BayesDel_noAF here against RyR1 variants is inflated by circularity, having been trained directly on many of the known variants in our test set. We also note the strong ancestry bias recently observed by BayesDel_noAF [47], further supporting the role of circularity.

**Figure 2:**
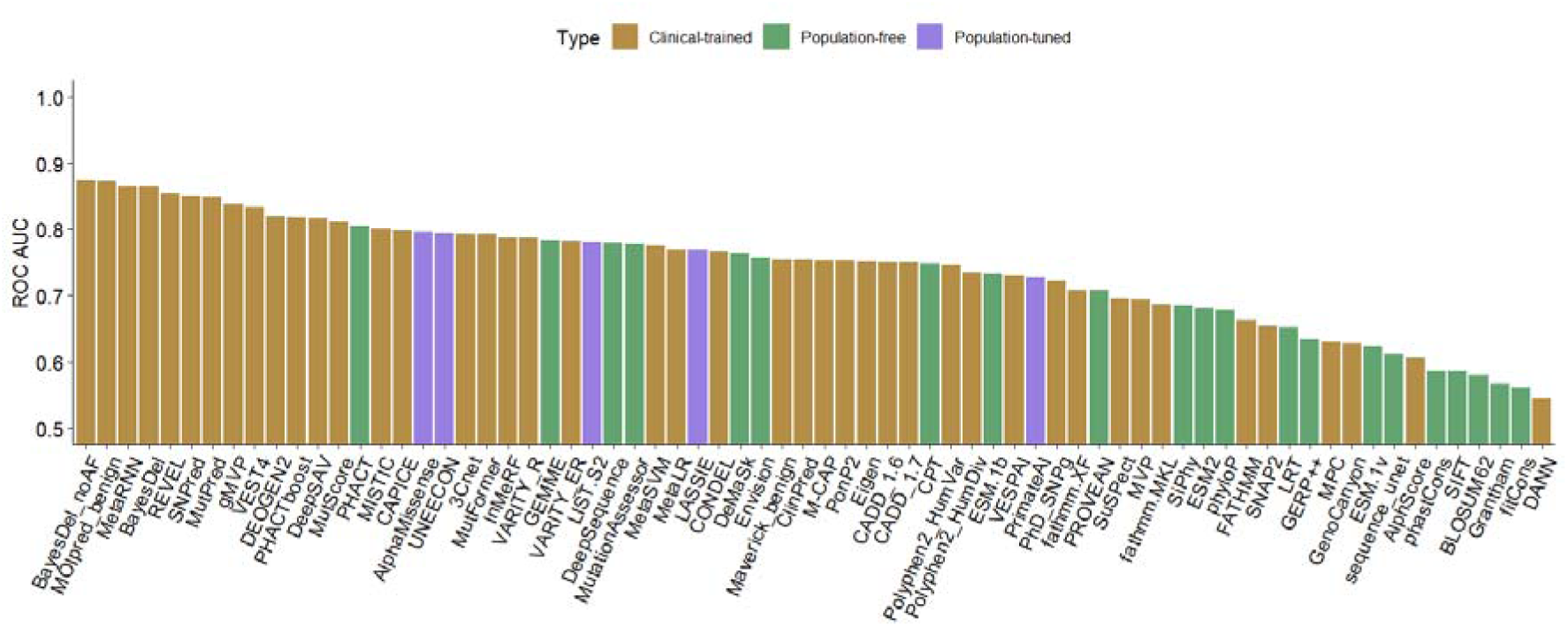
Performance of VEPs for the discrimination between pathogenic and putatively benign RyR1 variants. Bar plot showing the performance using ROC AUC (area under the receiver operating characteristic curve) for all the VEPs used in this study when evaluating pathogenicity of the variant dataset. VEPs are coloured by class: clinical-trained (brown), population-free (green) and population-tuned (violet).

Among the population-free VEPs, the highest ROC AUC is achieved by PHACT [49] (ROC AUC: 0.805). Interestingly, this method did not stand out in the recent DMS-based benchmark, ranking 41^st^ out of 97 tested VEPs[42]. Therefore, it is possible its performance here represents an element of random chance. Alternatively, we could speculate that perhaps PHACT uses high quality sequence alignments for RyR1, which may be difficult for other methods to align effectively due to the gene’s large size. Interestingly, the top performing VEP in that study, CPT-1, which combines sequence alignments, language models and protein structure [50], obtains a ROC AUC of only 0.748. AlphaMissense, a population-tuned predictor that ranked 2^nd^ (behind CPT-1) in the recent benchmark, achieved a ROC AUC of 0.796. While it avoids clinical label training, its reliance on population data introduces some risk of mild circularity.

Overall, the performance reported here as measured by ROC AUC values, appears to be fairly weak. In part, this may be related to the previous observation that gain-of-function missense variants are poorly predicted by current VEPs. Notably, the performance here is on a very similar level as reported for gain-of-function variants in that study. For example, DeepSequence [26] was found to have an average ROC AUC of 0.86 for genes with loss-of-function disease mechanisms and 0.80 for genes with gain-of-function disease mechanisms when discriminating between pathogenic ClinVar and putatively benign gnomAD missense variants mechanisms [20]. This is slightly lower but similar to the ROC AUC of 0.780 observed for DeepSequence here using similar datasets.

We also considered whether predictive performance might be different for variants associated with different dominant disease phenotypes. When performing assessments on a per-phenotype basis (*e.g.,* discrimination between MH and gnomAD missense variants), the relative performance of most VEPs changes very little, with the same few clinical-trained methods ranking best across all three groups (**Figure S1**). However, it does appear that MH are less well predicted than those associated with CCD and RRD. For example, CPT has a ROC AUC of 0.678 on the MH subset, compared to 0.822 and 0.798 for the CCD and RRD subsets. Since MH is an exposure-dependent genetic disorder, it is likely underdiagnosed relative to CCD and RRD, and its associated variants may exert weaker effects on fitness. As a result, they may be more difficult for computational predictors to identify.

Ideally, to mitigate circularity, one would exclude any variants used in VEP training from evaluation datasets. However, in practice this is not possible for most methods included here. Many clinical-trained VEPs, particularly metapredictors which themselves utilise scores from other VEPs, do not make their exact training sets or development timelines publicly available. Moreover, reconstructing which variants were accessible to predictors at the time of training is complicated by the lack of detailed version histories for most VEPs and the difficulty of aligning their training timelines with ClinVar submission dates. This opacity is itself a contributor to circularity and limits the feasibility of rigorous de-overlapped evaluations. As such, we adopt a gene-centric approach and interpret results in light of known biases, highlighting the superior performance of population-free VEPs in DMS benchmarks as a more reliable indicator of generalisability.

### Protein structural analysis of Ryr1 missense variants

Most VEPs in this study utilise only protein sequence information, with no inclusion of three-dimensional protein structures [51]. Given the limitations of current VEPs for the identification of pathogenic missense variants in RyR1, we wondered whether consideration of protein structural information could help us to improve upon this.

Previously, it has been shown that predicted changes in protein stability can be useful for the identification of pathogenic missense variants in some proteins, particularly those associated with recessive disorders and haploinsufficiency [20, 52]. Therefore, we first assessed whether the predicted effects of missense variants on protein stability showed any difference between pathogenic and putatively benign variants, or between the different phenotypic classes. Notably, |ΔΔG| showed essentially no ability to discriminate between pathogenic and putatively benign RyR1 missense variants, with a ROC AUC value of 0.557. The only significant differences were between putatively benign variants and RRD variants (*P* = 7.2 × 10^-5^, Wilcoxon rank-sum test), as well as MH and RRD (*P* = 0.02, Wilcoxon rank-sum test) (**Figure S2**). The MH variants had a lower |ΔΔG| than the more heterogeneous RRD variants, which is consistent with the fact that this phenotypic category’s mechanism of disease is gain of function, which tend to not be structurally disruptive. In contrast, the heterogeneous RRD group is more likely to include some loss of function variants, which are expected to have higher ΔΔG values than benign and therefore likely to be structurally disruptive. However, our results suggest that protein destabilisation or disruption of inter-subunit interactions is not a major contributor to any class of RyR1 pathogenic variants, especially given its poor performance in variant classification.

In previous work from our group, it was found that pathogenic gain-of-function and dominant-negative variants show a strong tendency to cluster in three-dimensional space; it was therefore suggested that this clustering could potentially be used to improve the predictions of mutations associated with such non-loss-of-function mechanisms [20]. Since previous work suggests that most dominant RyR1 variants are associated with gain-of-function effects [6], which is further supported by our ΔΔG analysis, we next decided to assess the level of mutation clustering in RyR1.

Some evidence for clustering of pathogenic RyR1 missense variants is immediately apparent when inspecting the one-dimensional sequence-level plot in **Figure 1**, most notably with the high density of variants near the C terminus, particularly those associated with CCD. When we plot the positions of these mutations onto the structure of the RyR1 tetramer (**Figure 3**), the clustering around the C-terminal pore region becomes even clearer. Additionally, we can see that distinct placements emerge gathered around the N terminus, the middle of the sequence, and the C terminus. We note the enrichments of these variants in the MIR (protein mannosyltransferase, IP3R, and RyR) domains at the N terminus of the sequence [53]. Specifically, based on these clustering analyses, MH variants mostly clustered in the Nsol, Bsol, pVSD, pore, and CTD domains, CCD variants clustered in the pVSD, pore, and CTD domains, RRD variants clustered in the NTD, Nsol, pore, and CTD, while recessive variants were spread across the structure [5, 54, 55]. Interestingly, we see that CCD variants cluster most strongly in the C terminus at the lower extreme of the protein complex, which corresponds to the channel and activation (CAC) domains. This is mechanistically interesting, as CCD is thought to occur due to incomplete closure of the ion channel, thus resulting in permanent leakage of calcium. The MH variants are also shown to cluster around the CTD, which has been shown to harbour a Ca^2+^ binding site, as well as an allosteric tryptophan residue (W4716), which is mechanistically interesting as in malignant hyperthermia, as hypersensitivity to Ca^2+^ activation is one of its causative mechanisms [56–58].

**Figure 3:**
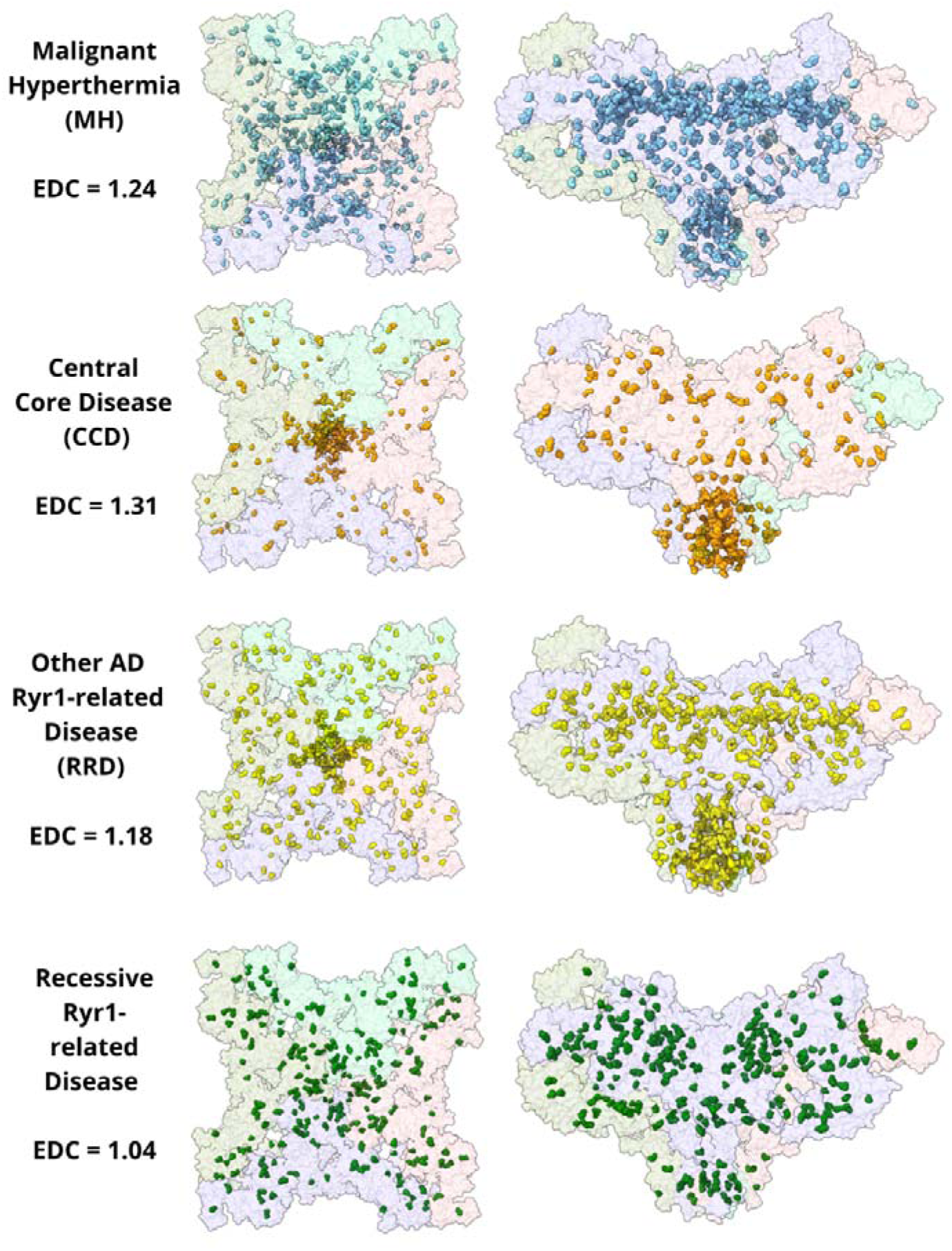
Clustering of pathogenic missense variants within the tetrameric structure of RyR1. The structure of rabbit RyR1 (PDB ID: 7M6A) is used, variant positions shifted with respect to the human sequence. Pathogenic missense variants are highlighted and coloured (blue: MH, malignant hyperthermia; orange: CCD, central core disease; yellow: RRD, other AD RYR1-related diseases, green: AR RYR1-related diseases (recessive). These images show similar patterns as in Figure 1, with missense variants appearing across the sequence in the top and side views.

To quantify the degree of spatial clustering of pathogenic variants, we used the recently introduced Extent of Disease Clustering (EDC) metric [20]. EDC values close to one indicate little-to-no spatial clustering of the sites of pathogenic mutations, *i.e.,* the sites of disease variants are essentially randomly dispersed throughout the protein. In contrast, EDC values greater than one indicate that the pathogenic mutations are clustered in three-dimensional space. Previously, the mean EDC value for pathogenic missense variants in autosomal recessive disease genes was 1.02, indicating very little tendency to cluster [20]. In contrast, gain-of-function and dominant-negative missense variants from autosomal dominant disease genes both had mean EDC values of 1.27, demonstrating their strong tendency to cluster together.

For RyR1, when we consider all pathogenic missense variants collectively, we obtain an EDC value of 1.16, indicative of moderate clustering and consistent with a mixture of gain- and loss-of-function mechanisms across phenotypic categories. When broken down by disease group, dominant variants show notably stronger clustering, with EDC values of 1.31 for CCD, 1.24 for MH, and 1.18 for RRD. In contrast, recessive variants yield an EDC of 1.04, indicative of minimal clustering and consistent with classical loss-of-function mechanisms [59]. These findings support the view that most dominant RyR1-related phenotypes are associated with gain-of-function effects, with CCD variants showing the most pronounced spatial clustering. The differences in EDC between phenotypic groups may reflect underlying variation in the strength or specificity of the gain-of-function mechanism involved.

### Using spatial clustering as a predictor of pathogenicity

Given the strong clustering observed for dominant RyR1 missense variants, we next explored whether this information could be used to improve the identification of pathogenic variants. Building upon the EDC metric, we introduced a simple spatial clustering feature based on the average distance between a given RyR1 amino acid residue and the *K* nearest other residues that are sites of known pathogenic variants, considering the positions of alpha-carbons in the structure (PDB ID: 7M6A), which we call Spatial Proximity to Disease Variants (SPDV). Since pathogenic variants tend to cluster in three-dimensional space, we reasoned that residues in close proximity to these known sites are more likely to contribute to similar structural or functional disruptions and thus may have a higher likelihood of pathogenicity when mutated. Previous studies have shown that spatial clustering of mutations can help identify functionally important regions or potential driver mutations, particularly in cancer [31, 60], but this has rarely been applied to rare disease variant classification in a clinically interpretable framework [61].

Importantly, while SPDV is based directly on the positions of known pathogenic variants, there is no potential for circularity in our evaluation of its performance. When evaluating the potential pathogenicity of a given mutation at a residue, the calculation only considers the unique locations of known pathogenic mutations at other residues, *i.e.,* the scoring of a known pathogenic mutation would not be influenced by the fact that it is known to be pathogenic.

In **Figure S3**, we test SPDV for the discrimination between pathogenic and putatively benign missense variants for a range of *K* values. We find the metric is relatively robust to the choice of *K*, but the top performance is with *K*=4 (SPDV_4), which achieves a ROC AUC of 0.762. This puts it slightly worse than the top population-free VEPs, but still demonstrates reasonable performance given the orthogonal strategy. An analogous 1D model calculated using distance in primary sequence showed worse performance (ROC AUC: 0.728), demonstrating the importance of considering protein structure. Finally, while SPDV used here is based only on the RyR1 subunit structure, considering only intramolecular distances, consideration of intermolecular distances from the full tetramer gave essentially equivalent results (not shown).

Given the overlapping but distinct patterns of clustering observed for the different dominant disorders, and the lack of clustering for recessive variants, we might expect differences in the performance of our spatial clustering model when applied to different mutation classes. Therefore, we repeated the analysis using each of the three dominant disease phenotypes separately **(Figure S4)**. Interestingly, predictive performance improves for the CCD model compared to treating all disease mutations collectively, with a top ROC AUC of 0.806 when using *K*=2. The MH model achieves very similar performance to SPDV calculated using the whole dataset, with a top ROC AUC of 0.760 (*K*=4).

In contrast, the RRD model performs modestly worse with a top ROC AUC of 0.713 (*K*=1). The relatively weaker performance of SPDV for RRD-associated variants likely reflects the underlying heterogeneity of this group. Unlike CCD, which is mechanistically homogeneous and strongly clustered at the C-terminal pore region, the RRD group includes a mix of dominant phenotypes with diverse clinical presentations and potentially distinct pathogenic mechanisms. In particular, this set may contain both gain- and loss-of-function variants, as suggested by the broader distribution of ΔΔG values observed for RRD compared to CCD and MH (Figure S2). Since SPDV is predicated on spatial clustering typical of gain-of-function or dominant-negative mechanisms, the inclusion of more diffuse or structurally destabilising variants within RRD naturally diminishes its predictive power.

Overall, it appears that mutation classes that exhibit a greater degree of clustering, as measured by EDC values, are also better predicted by our SPDV model, especially the CCD phenotype group which also has the highest EDC of 1.31. All the SPDV outputs from our 14 models are available in **Table S4**. The strong performance of SPDV for CCD variants, driven by tight clustering near the C-terminal pore, suggests that this approach may be particularly well suited to ion channels and other macromolecular complexes where pathogenic variants localise to functionally critical structural domains. Future application to other large ion channels or receptors with known clustering, such as *SCN1A* [62] or *CACNA1A* [63], could help assess how widely structure-based proximity metrics can support variant interpretation.

### Clinical classification of RyR1 missense variants

To assess the practical utility of both our SPDV feature and VEPs in the context of clinical variant interpretation, we classified missense variants in *RYR1* according to ACMG/AMP guidelines, specifically focusing on the PP3 and BP4 criteria [64, 65]. While previous efforts applied genome-wide calibration of VEPs for this purpose [66], we employed a gene-specific approach, utilising our recently released *acmgscaler* tool that utilises a robust kernel density estimation method [32]. This gene-specific approach is greatly facilitated by the large number of *RYR1* clinically classified variants available for calibration. While the ROC AUC analyses above used less stringent criteria for variant inclusion, here, due to the potentially direct clinical applicability of this calibration, we use a stricter truth set comprising expert-curated (likely) pathogenic and (likely) benign missense variants from the VCEP [34].

For each variant, we estimated the positive likelihood ratio for each variant. After assigning evidence strengths to each variant based on the score-specific likelihood ratios, we focused on the SPDV instance SPDV_4 based on its previous performance. To avoid potential biases inherent in the use of clinical-trained VEPs, as discussed previously, we compared SPDV_4 to the highest-performing population-free VEP by ROC AUC, PHACT. Full classification results and evidence thresholds are provided in **Table S5**, offering a resource for future variant interpretation efforts. The calibrations for the two approaches are shown in **Figure 4A**. While PHACT reaches a higher maximum evidence level for pathogenicity (PP3_strong) than SPDV_4 (PP3_moderate), SPDV_4 achieves a much stronger benign classification (BP4_strong) than PHACT (BP4_supporting).

**Figure 4:**
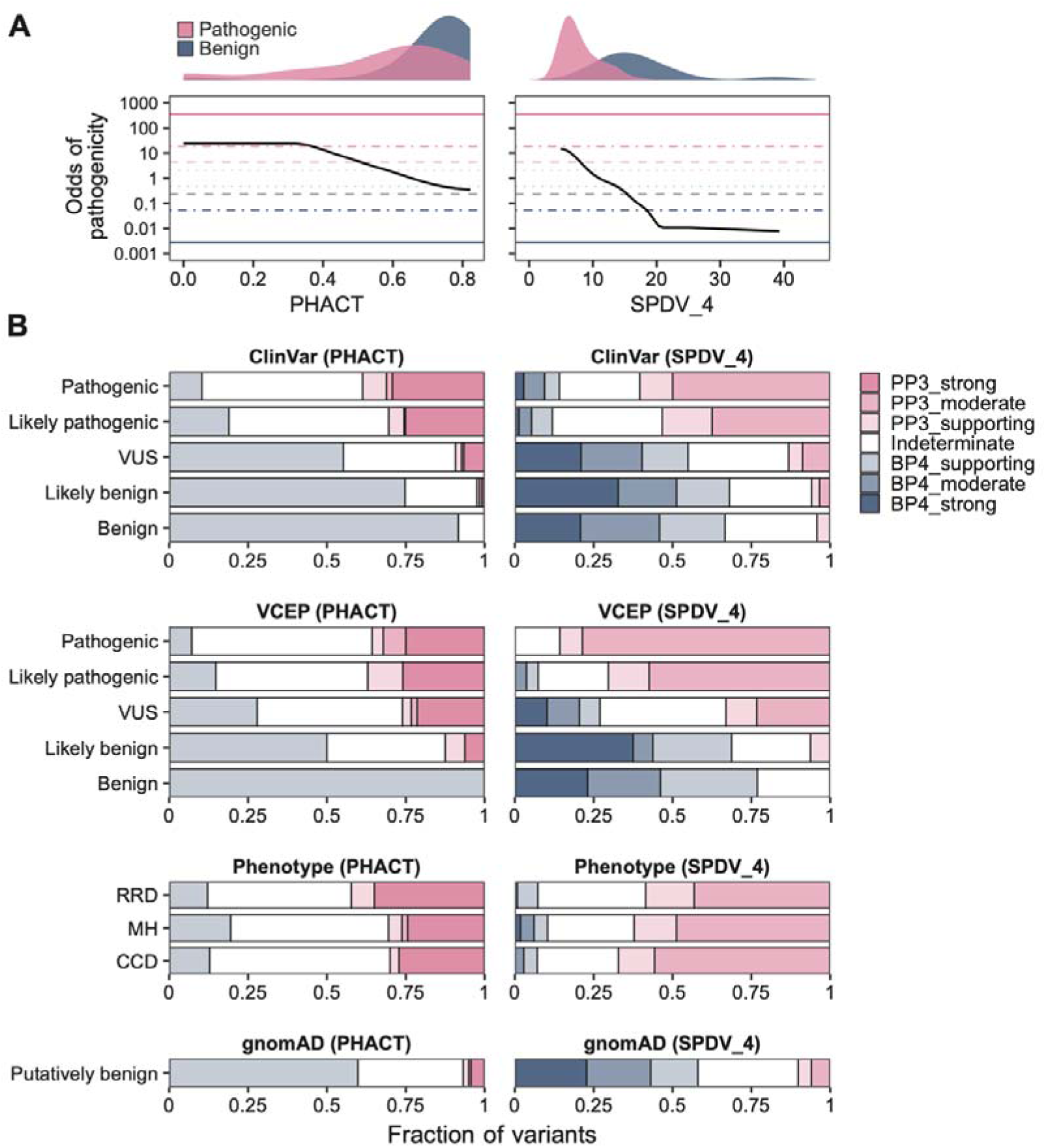
Comparison of ACMG/AMP variant classification outcomes for RyR1 missense variants between PHACT and SPDV_4. **(A)** PHACT and SPDV_4 plotted against their odds of pathogenicity values. Horizontal lines show the odds of pathogenicity thresholds corresponding to the PP3/BP4 evidence levels, which were derived from the very strong threshold for pathogenicity based on a 10% prior. Density plots on the top show the distribution of benign (ClinVar B/LB) and pathogenic (ClinVar P/LP) variants. **(B)** Composition of PP3/BP4 evidence classes reached by PHACT and SPDV_4 in the following missense variant categories: gnomAD v4.1, ClinVar, Variant Curation Expert Panel (VCEP), and Phenotype, which includes the dominant phenotypes RRD, MH and CCD.

When applied to pathogenic or likely pathogenic variants from ClinVar or the VCEP, an interesting pattern emerges (**Figure 4B)**. While PHACT is able to provide strong evidence of pathogenicity to a subset of disease-associated variants (∼25%), SPDV_4 is able to provide moderate or supporting evidence to a much larger proportion (∼50-80%). The trend when applied to benign or likely benign variants is the opposite: PHACT is able to provide supporting evidence to a greater proportion (∼50-100%), but SPDV_4 can provide moderate or strong evidence for many (∼40-50%). These findings suggest the two methods are complementary: SPDV_4 provides broader but weaker pathogenic support, while PHACT assigns stronger evidence to fewer variants, and vice versa for benign evidence.

When considering VUS from ClinVar, most receive at least supporting evidence of benignity using either method, with only a small proportion receiving evidence of pathogenicity. This suggests that most reported VUS in ClinVar are probably actually benign. In contrast, the VCEP variants are much more evenly split, with ∼25% receiving evidence of pathogenicity and ∼25% receiving evidence of benignity with both methods. This suggests that the VUS identified by the VCEP experts are much more likely to actually be pathogenic, in contrast to ClinVar, where large numbers of population variants are being classified as VUS. It also highlights the potential power of using these computational tools for prioritising *RYR1* VUS.

When applied to disease-associated variants associated with different phenotypes, it is interesting to note that PHACT provides the highest proportion of pathogenic evidence for RRD variants, whereas SPDV_4 is most effective for CCD variants, which display the highest level of spatial clustering. This is similar to what was observed in the ROC AUC analysis (**Figures S1 and S4**).

Finally, we assessed classifications for putatively benign gnomAD variants. Most (∼60%) of these variants received evidence of benignity using either method, with only ∼5-10% receiving evidence of pathogenicity. This likely reflects the presence of a small subset of pathogenic variants in the population, due to factors such as recessive inheritance or the latent, exposure-dependent nature of MH.

## Conclusions

Although VEPs are valuable tools for investigating VUSs, they can suffer from biases that can impact their use in clinical settings. In the case of RyR1, the lack of adequate computational VUS interpretation places burden both on clinicians and prospective patients with uncertain clinical diagnoses for an illness in the RyR1 disease spectrum. Due to the phenotypic variation for disease in RyR1 and its extremely long primary sequence, missense variant classification is challenging. In this study, we performed an evaluation of 70 VEPs on RyR1 pathogenic and putatively benign missense variants. Our results suggest that existing computational methods perform poorly in RyR1 variant classification, across all dominant clinical phenotypes.

Previous work from our group investigated the phenomenon of disease variant clustering in protein structures using the EDC metric [20]. Building on this, we have here introduced a novel feature based on this phenomenon using mean minimum distances to pathogenic variants in 3D space (SPDV) that resulted in comparable variant classification compared to current VEPs, with the highest performing iteration achieving 0.762 ROC AUC (SPDV_4). Using our clustering metric and structural visualisations of the variants in the RyR1 homotetramer, we demonstrated strong clustering of disease variants in 1D and 3D space. Most notably, CCD variants are extremely clustered at the C terminus of the protein as is shown in the literature [67, 68]. These changes are also reflected in the different values for the EDC metric that we calculated for the MH, CCD, RRD, and recessive classes. The MH and CCD variant classes exhibited EDC metrics that were very similar to those of autosomal dominant disease genes associated with dominant-negative and gain-of-function mechanisms [20]. These analyses suggest that distance to pathogenic variants represents a viable and computationally simple strategy for predicting variant pathogenicity. We expect that this could ultimately provide better and more intuitive interpretations of pathogenicity in future analyses of other proteins by avoiding the biases of previous clinical-trained VEPs.

In this study, we classified missense variants in RyR1 from ClinVar using a gene-specific application of odds of pathogenicity thresholds and ACMG/AMP PP3/BP4 evidence levels. Among the population-free VEPs evaluated, PHACT exhibited the highest discriminatory power in distinguishing pathogenic from putatively benign variants. However, compared to SPDV_4, PHACT assigned supporting evidence to nearly twice as many variants in gnomAD, an unexpectedly high proportion given that truly pathogenic variants should be rare in population databases. While PHACT achieved higher ROC AUC values, its performance was limited by substantial score overlap between pathogenic and benign variants, which constrained its ability to assign higher evidence levels for pathogenicity. This highlights the limitations of relying solely on ROC AUC as a measure of clinical utility. To more effectively assess the relevance of VEPs in variant classification, future approaches should account for the full score distribution and the distinction between benign and pathogenic variants across the entire range of predictions.

The computational simplicity of SPDV, coupled with performance can rival or surpass population-free VEPs in RyR1, underscores its value as a structure-based approach for variant prioritisation. It is particularly useful for identifying gain-of-function variants, providing an orthogonal strategy that complements existing sequence-base predictors like PHACT. Given this complementarity, it may be reasonable to select the method (*i.e.* between SPDV and a sequence-based VEP like PHACT) that gives the strongest level of evidence, so long as the approaches are truly independent. Crucially, one should avoid cherry-picking among sequence-based VEPs to maximise support for a variant, as this can lead to overstatement of clinical evidence [66]. However, given their independent nature (assuming the VEP does not utilise any analogous distance metric), this approach should not introduce such bias. Ultimately, the question of how best to integrate orthogonal sources of evidence, like evolutionary constraint and structural proximity, remains unresolved, mirroring similar challenges in integrating multiplexed experimental data with computational predictions [69]. Future extensions of SPDV could also incorporate weighting schemes or additional evidence sources to further refine its predictive resolution, particularly in contexts where pathogenic variants exhibit less pronounced clustering.

More broadly, our results illustrate how gene-specific structural insights can inform practical tools for variant interpretation, particularly in proteins where current predictors underperform. Spatial proximity metrics, while conceptually simple, offer interpretable and flexible improvements that integrate well into existing variant classification frameworks. These findings point to a promising direction for expanding structure-informed approaches across other genetically and structurally complex disease genes.

## Data availability

All datasets associated with this study, including variants and associated variant effect scores, are provided as Supporting Information.

## Acknowledgements

We thank Karolina Jackowska for help with compiling the dataset of pathogenic variants, Benjamin Livesey for assistance with the variant effect predictor calculations and Elaine Fletcher for helpful discussions.

## Supporting Information

**Supporting Information 1.** Table S1: Pairwise sequence alignment between human RyR1 and the rabbit PDB structure 7M6A.

**Supporting Information 2.** Table S2: Pathogenic missense variants and their sources. This table includes all the variants gathered from the various databases and literature searches we used to gather as many RYR1 pathogenic missense variants as possible. Here we have provided the ‘Missense Variants’ tab, which includes the variants and their sources, as well as the ‘Literature’ tab, which contains the paper references for the relevant variants.

**Supporting Information 3.** Table S3: RYR1 complete dataset and ROC AUC evaluations using the full dataset and phenotype-specific sets. This table includes the pathogenic missense variants together with the putatively benign variants from gnomAD, along with their associated VEP scores. Here we have provided the ‘Complete Dataset’ tab, which includes all the aforementioned data, as well as the ‘Complete Dataset’, ‘AUROC’ and ‘By phenotype’, ‘AUROC’ tabs, which contain the results of the VEP ROC AUC evaluations for these sets.

**Supporting Information 4.** Table S4: SPDV values for all residues in RYR1 and ROC AUC evaluations using the full dataset and phenotype-specific sets. This table includes all the SPDV values calculated using K=1-10,20,30,40, and 50 for all the residues in RYR1. Here we have provided the ‘SPDV 1-50’ tab, which includes all the aforementioned data, as well as the ‘SPDV Complete Dataset’, ‘AUROC’ and ‘SPDV By phenotype’, ‘AUROC tabs’, which contain the results of the SPDV ROC AUC evaluations for these sets.

**Supporting Information 5.** Table S5: ACMG values and ACMG thresholds for VEPs. This contains the score intervals for the given ACMG/AMP PP3/BP4 evidence strengths.

**Figure S1:**
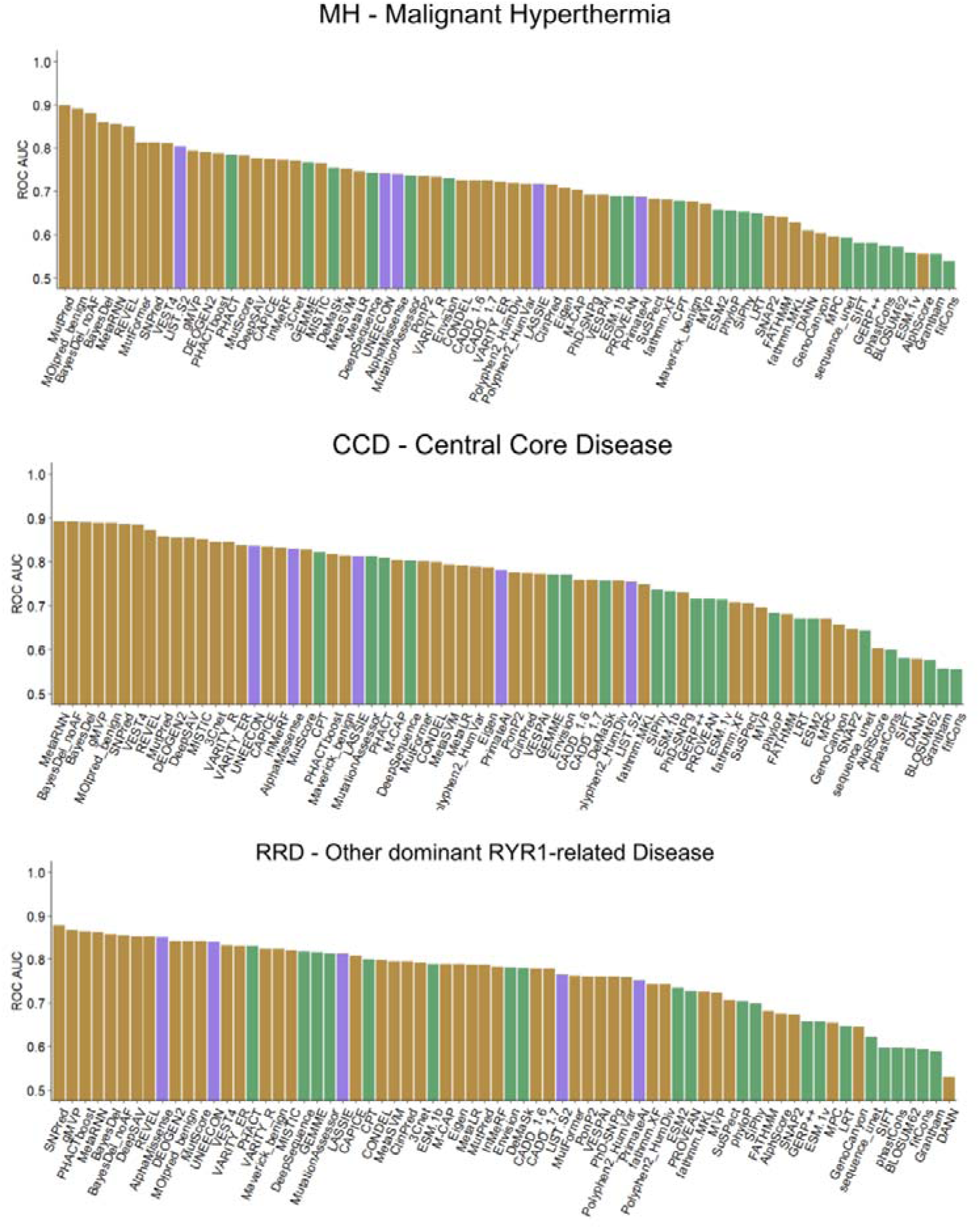
VEP performance for the identification of pathogenic RyR1 variants by phenotypic group. Bar plot showing the performance using ROC AUC (area under the receiver operating characteristic curve) for all the VEPs used in this study when evaluating pathogenicity of variants by the MH (malignant hyperthermia), CCD (central core disease), and RRD (other dominant RYR1-related disease) groups when compared to putatively benign variants. VEPs are coloured by class: clinical-trained (brown), population-free (green) and population-tuned (violet).

**Figure S2:**
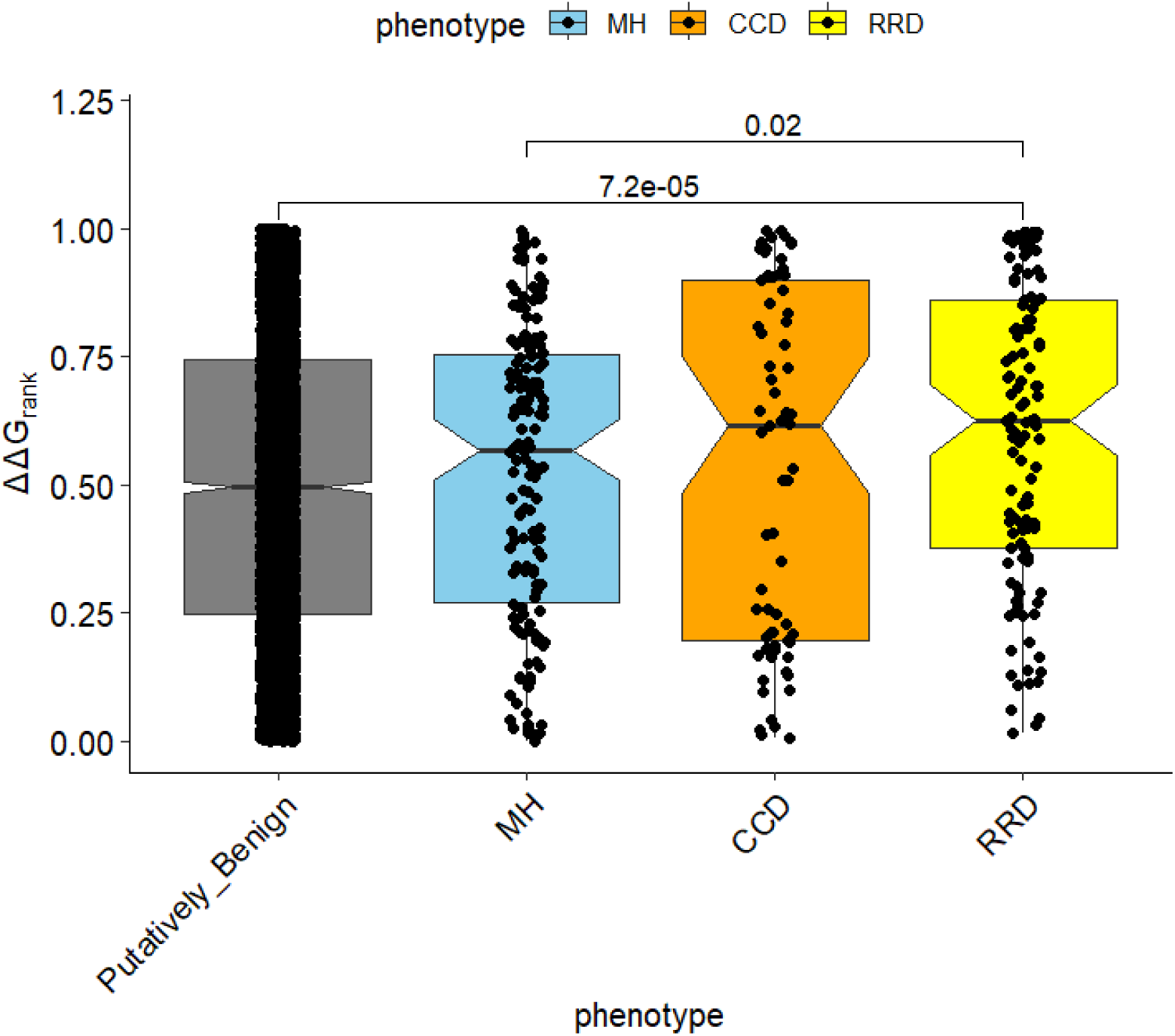
Comparison of ΔΔG_rank_ values from missense variants associated with different RYR1 phenotypic groups. ΔΔG_rank_ is a recently introduced rank-normalised representation of |ΔΔG| that improves visual comparisons (10.1016/j.celrep.2024.114905).

**Figure S3:**
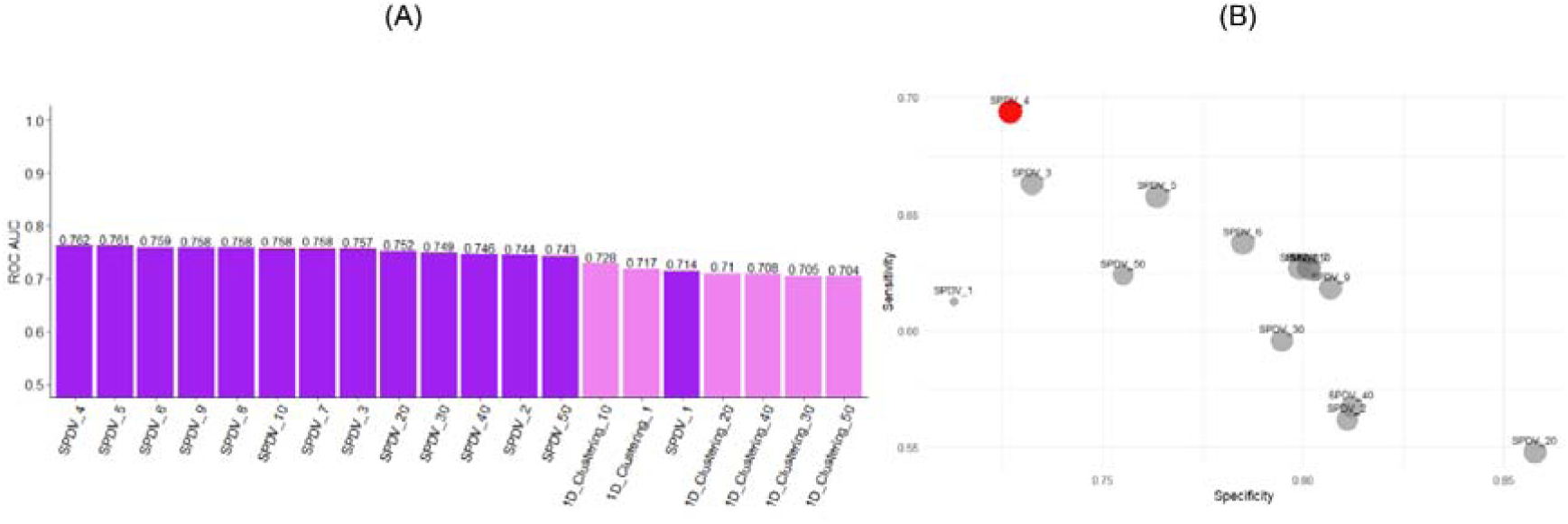
SPDV_4 is the top performing SPDV instance. (A) ROC AUC performance of different SPDV instances calculated using the 3D protein structure and the mean distance to the K nearest pathogenic variants, where K = 1–10, 20, 30, 40, and 50. While performance is relatively consistent across values of K, SPDV_4 achieves the highest AUC. (B) Sensitivity versus specificity for the same set of SPDV instances, evaluated at the optimal threshold determined by the Youden index (J statistic). SPDV_4 shows the best balance between sensitivity and specificity, further supporting its selection for downstream analysis.

**Figure S4:**
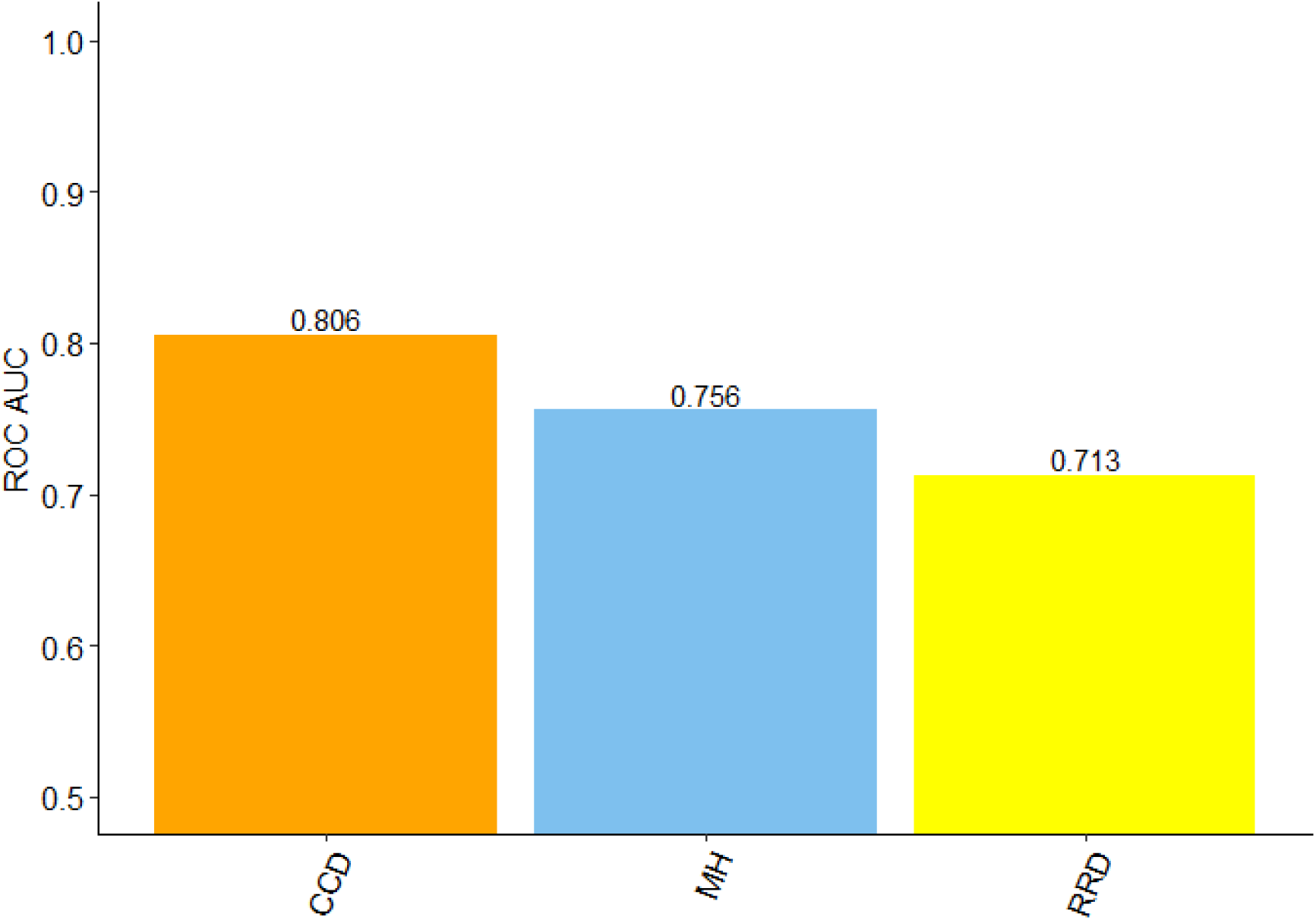
Performance of the top performing SPDV instances by phenotypic group. Predictive performance improves for the CCD model compared to treating all disease mutations collectively, with a top ROC AUC of 0.806 (*K*=2). The MH model achieves very similar performance to SPDV calculated using the whole dataset, with a top ROC AUC of 0.760 (*K*=4). The RRD model performs slightly worse with a top ROC AUC of 0.713 (*K*=1).

